# Automated quantitative evaluation of thymic involution and hyperplasia on plain chest CT

**DOI:** 10.1101/2023.11.13.23298440

**Authors:** Yuki T. Okamura, Katsuhiro Endo, Akira Toriihara, Issei Fukuda, Jun Isogai, Yasunori Sato, Kenji Yasuoka, Shin-Ichiro Kagami

## Abstract

**Objective:** To establish an automatic method to quantify thymic involution and hyperplasia based on plain chest computed tomography (CT).

**Methods:** We defined the thymic region for quantification (TRQ) as the target region. We manually segmented the TRQ in 135 CT studies, followed by construction of segmentation neural network (NN) models based on the data. We developed the estimator of thymic volume (ETV), a measure of the thymic tissue volume in the segmented TRQ. The Hounsfield unit (HU) value and volume of the TRQ were measured, and the ETV was calculated in each CT study from 853 healthy subjects. We investigated how these measures were related to the age and sex using quantile additive regression models. We defined the ETV z-score, an age- and sex-adjusted version of ETV, to distinguish between subjects with thymic hyperplasia (18 cases) and healthy subjects. A receiver operating characteristic (ROC) curve analysis was conducted.

**Results:** A significant correlation between the NN-segmented and manually segmented TRQ was seen for both the HU value and volume of the TRQ (*r* = 0.996 and *r* = 0.986 respectively). The ETV could detect age-related decline in the thymic tissue volume (*p* < 0.001). No statistically significant difference was detected between male and female subjects (*p* = 0.19). The ETV was significantly higher in the thymic hyperplasia group as compared with that in the healthy control group (*p* < 0.001). The ETV z-score could distinguish between subjects with thymic hyperplasia and healthy subjects, with the ROC curve analysis revealing an area under the curve (AUC) of 0.88 (95% CI: 0.75-1.0).

**Conclusion:** Our method enabled robust quantification of thymic involution and hyperplasia. The results were consistent with the trends found in previous studies.

**Clinical Relevance Statement:** Our method allows reliable and automatic measurement of thymic involution and hyperplasia on CT images. This may aid in the early detection and monitoring of pathologies related to the thymus, including autoimmune diseases.

**Key Points:**

- We defined the thymic region for quantification (TRQ) to fully automate the evaluation of thymic involution and hyperplasia. The neural networks could identify the TRQ with sufficient accuracy.
- We developed the estimator of thymic volume (ETV) to quantify the thymic tissue in the TRQ. ETV captured age-related thymic involution and thymic hyperplasia.
- The ETV could prove useful in the management of pathologies associated with involution or hyperplasia of the thymus.

## Introduction

Thymus is one of the most important organs comprising the human immune system, where T cells mature, and harmful self-reactive T cells are eliminated [1]. In children, the thymus is visualized on chest computed tomography (CT) as a soft tissue density in the anterior mediastinum. As one ages, it is gradually replaced by fat density reflecting age-related involution. Finally, some residual thymic tissue remains as scattered spots in the mediastinal adipose tissue [1, 2]. Thymic involution and loss of thymic function are associated with a higher mortality and a higher risk of cancers and autoimmune diseases in human adults [3–5].

Thymic hyperplasia is observed when aberrant cell proliferation occurs due to various pathologies such as infections, autoimmune diseases, steroid usage, and serious stress. Identification of thymic hyperplasia could lead to an accurate diagnosis of these underlying pathologies [6, 7]. In clinical practice, visual inspection or measurement of the thymus thickness is often used for radiological diagnosis of thymic hyperplasia [8, 9]. Detection of thymic hyperplasia is not easy, since the normal thymic tissue volume differs greatly by age. As a result, the early stages of thymic hyperplasia can often be overlooked.

Previous studies have used visual scoring or Hounsfield unit (HU) value inside the region of interest (ROI) for roughly estimating the thymic tissue volume [2, 10–13]. However, these methods are only semi-quantitative and cannot directly estimate the quantity of thymic tissue. They are also too time-consuming for daily clinical use or large-scale clinical studies. One of the most effective solutions for these problems is a neural network (NN)-based system to automatically segment and quantitatively evaluate the thymic region. Such tasks present challenges, because of the small volume and indistinct borders of the thymic region.

The purpose of this study was to develop a method to automatically identify the thymic region on chest CT images and to quantify the thymic tissue volume to evaluate thymic involution and hyperplasia.

## Materials and methods

### Study design and datasets

This retrospective study was conducted with the approval of the institutional review boards of Asahi General Hospital and Keio University. CT images and other clinical data were collected from April 2006 to March 2022. Three datasets were prepared to develop and evaluate the system.

The development dataset consisted of CT studies from 135 subjects, that were used for training, validation, and testing of the segmentation NN. The chest CT images were selected randomly and were derived from subjects with a wide range of demographic backgrounds, in order to achieve a high generalizing performance. The inclusion criteria for the images were as follows: (1) axial CT images without contrast media, (2) slice thickness of 1-3 mm, (3) no lesions in the mediastinum, such as thymoma, lymphadenopathy, or thoracic aortic aneurysm, (4) no pulmonary or pleural lesions adjacent to the frontal or upper mediastinum, and (5) no history of sternotomy. The same inclusion criteria were also applied for the following datasets.

The healthy dataset consisted of CT studies from 853 subjects who underwent CT for investigation of abnormal chest X-ray findings. This dataset was used for the evaluation of age-related thymic involution. The CT studies revealed negative or benign findings in all the subjects. Subjects with signs of acute illness, a history of systemic corticosteroid or anabolic steroid use, a history of thymus-related diseases, or a history of systemic diseases associated with major chronic inflammation were excluded.

The thymic hyperplasia dataset consisted of CT studies from 18 subjects, which radiologists had interpreted as showing thymic hyperplasia.

Every dataset included one study per subject. All CT images were acquired using one of the multi-detector CT scanners at our hospital. See the Supplementary Methods for more details.

### Automatic segmentation of the thymic region for quantification (TRQ)

Since accurate and robust segmentation is needed for appropriate evaluation of the thymic region, we defined the thymic region for quantification (TRQ) as a three-dimensional region in the anterior mediastinum (Fig. 2) bordered by the following structures:

Superior border: the level at which the left brachiocephalic vein passes in front of the aorta or the brachiocephalic artery.

Inferior border: the level at which the anterior wall of the aorta and the main pulmonary artery line up right-to-left.

In each study in the development dataset, manual segmentation of the TRQ was performed by consensus between one physician and one radiologist, using the ITK-SNAP software [14]. First, the airways were labeled using the snake tool of ITK-SNAP, with an upper threshold of −300 HU. Next, the TRQ was identified manually according to the abovementioned definition. The TRQ HU value histogram of the segmented TRQ for each image was visually inspected.

For automatic identification of the TRQ in CT images, NN models (DeepLabV3 NN with a ResNet-50 backbone) [15] were trained and validated (Fig. S1). Each axial slice in a CT study was used as an input to the NN. The NN models were implemented in Python (v 3.9), using PyTorch (v 1.10). The development dataset was split randomly into six subsets. One subset was kept as the test set, while the remaining five subsets (training set) were used for NN training and five-fold cross validation. Five NN models were generated as a result of this five-fold cross validation step. See the Supplementary Methods for more details.

### Evaluation of the segmented TRQ

The performance of the segmentation NN models was assessed using the test set. Each CT study in the test set was processed by the five NN models generated in the five-fold cross validation step, and the five segmentation results were processed separately. We comparatively evaluated the segmentation accuracy between automatic segmentation and manual segmentation by determining the Dice similarity coefficient (DSC) [16]. We created yy plots and Bland-Altman plots to compare the HU values and volumes between the NN-segmented and manually segmented TRQ. We used the mode value as the representative TRQ HU value. If the mode value was impossible to calculate, the segmentation result was discarded as invalid. See the Supplementary Methods for more details on estimation of the TRQ HU value. The median of the five values calculated from the five segmentation results was used as the representative value for each CT study, for DSC, TRQ HU value and volume. We also calculated Pearson correlation coefficients and intraclass correlation coefficients (ICC; two-way random, single measures, absolute agreement) for the TRQ HU values and volumes. The ICC was calculated using the psych package on R. The DSC obtained from the five-fold cross-validation is also reported.

In order to eliminate low-quality segmentation results, we attempted to conduct quality control after the TRQ segmentation. In the process of quality control, we needed to predict the segmentation quality using measures that are computable based on the five automatic segmentation results. To establish the criteria for this process, several measures including the HU value variance and mean pairwise Jensen-Shannon divergence (JS divergence) were calculated. Pairwise correlation analysis between the segmentation quality and these measures were performed in the test set. The TRQ volume estimation error, HU value estimation error, and DSC were used as indicators of the segmentation quality. We determined the quality control criteria based on the correlation analysis results, and all the images that did not satisfy the criteria were excluded from the subsequent analyses. See the Supplementary Methods for more details.

### Quantifying the thymic tissue volume

While the TRQ HU value reflects the thymic tissue volume, the HU value could also be affected by the volume of the adipose tissue contained in the TRQ. To evaluate the thymic region with adjustment for this effect, we developed the estimator of thymic volume (ETV), which is a statistical estimator of the thymic tissue volume. ETV is defined as

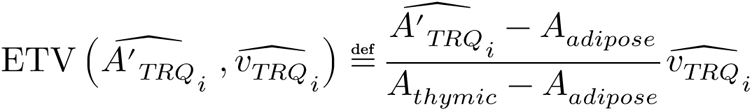

where *A*_*thymic*_ and *A*_*adipose*_ are predetermined constant HU values of thymic and adipose tissue, 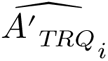 is the adjusted TRQ HU value, and 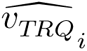 is the TRQ volume for CT study *i*. We examined the relationship between the TRQ volumes and HU values in the healthy dataset, to validate the theory of ETV. The derivation of the ETV is included in the Supplementary Methods. The theory of ETV is shown in Fig. S2.

To assess the capability of ETV for evaluating thymic involution and hyperplasia, images from the healthy dataset and thymic hyperplasia dataset were processed as shown in Fig. 1. Chest CT images were given as input to the five independent segmentation NN models generated in the five-fold cross validation step, to automatically identify the TRQ. The mode HU value and volume of the TRQ, as well as the ETV, were calculated for each of the five segmentation results. If any of the segmentation results were invalid among the five, the CT study was excluded from the analysis. Quality control was performed to remove low-quality segmentation results. Finally, the median value of the five calculated ETV values was considered as the representative ETV value.

**Fig. 1.**
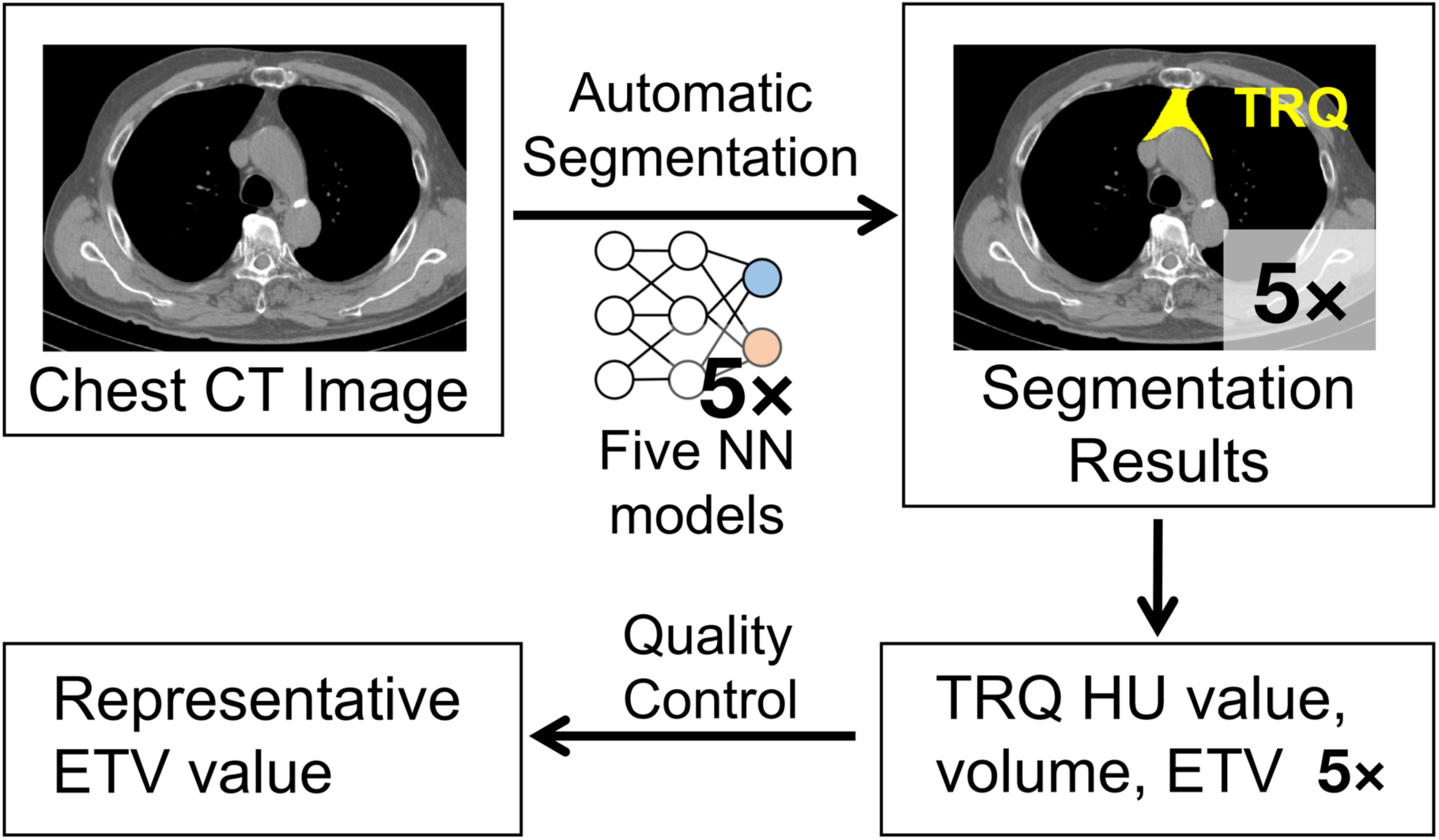
Workflow of the image processing. NN, neural network; TRQ, thymic region for quantification; HU, Hounsfield unit; ETV, estimator of thymic volume.

#### Measuring age-related changes of the thymic region

Age-related changes of the TRQ HU value, volume, and ETV were investigated in the healthy dataset. Differences in the values between male and female subjects were also assessed. To cope with the expected nonlinearity and unequal variances of the data, we chose the additive quantile regression model for the analyses (see the Supplementary Methods for more details).

#### Evaluation of thymic hyperplasia

To make the ETV value understandable in comparison with the healthy subjects, we introduced the ETV z-score, an age- and sex-adjusted version of the ETV. A detailed explanation of the ETV z-score is provided in the Supplementary Methods. Briefly, the ETV z-score shows how much larger or smaller the ETV in each subject is, in comparison with that in the age- and sex-matched population from the healthy dataset. An ETV z-score of zero is defined by a value equal to the median value in the matched population. In subjects with ETV values higher or lower than the median, the ETV z-score will be a positive or negative number, respectively. The ETV z-score was calculated for each subject of the thymic hyperplasia dataset.

One sample two tailed t test was performed to verify if the mean ETV z-score of the thymic hyperplasia dataset was higher than zero. A Receiver operating characteristic (ROC) curve analysis was conducted to determine the ability of the ETV z-score to differentiate subjects with thymic hyperplasia from healthy subjects. The cutoff value was determined based on Youden’s index.

### Statistical analysis

All the statistical analyses were performed using R (v4.2.1). *p* < 0.05 was considered as denoting statistical significance in all the analyses.

## Results

### Characteristics of the patients and images

The development dataset, healthy dataset, and thymic hyperplasia dataset consisted of CT studies from 135, 853, and 18 subjects, respectively. The demographic features of these datasets are shown in Table 1, Table 2, and Table 3.

**Table 1.** Demographic information of the development dataset.

|  | Development dataset (135 subjects) |  |
| --- | --- | --- |
|  | Training set<br>(113 subjects) | Test set<br>(22 subjects) |
| Age (years) |  |  |
| -19 | 12 | 3 |
| 20-29 | 14 | 3 |
| 30-39 | 8 | 2 |
| 40-49 | 8 | 1 |
| 50-59 | 11 | 3 |
| 60-69 | 27 | 2 |
| 70-79 | 21 | 7 |
| 80- | 12 | 1 |
| Sex, female | 50 (44 %) | 14 (64 %) |

**Table 2.** Demographic information of the healthy dataset.

| Healthy dataset (853 subjects) |  |
| --- | --- |
| Age (mean±SD) (years) | 53.6 ± 15.9 |
| Sex, female | 413 (48 %) |

SD: standard deviation.

**Table 3.** Demographic information of the thymic hyperplasia dataset.

| Thymic hyperplasia dataset (18 subjects) |  |
| --- | --- |
| Age (mean ± SD) (years) | 48.6 ± 14.1 |
| Sex, female | 11 (61 %) |
| Background pathology |  |
| Connective tissue disease<br>(Rheumatoid arthritis, Sjögren's<br>syndrome, Mixed connective tissue<br>disease) | 10 |
| Interstitial pneumonia | 5 |
| Lymphoma | 1 |
| Nephrosis | 1 |
| Bacterial pneumonia | 1 |

SD: standard deviation.

### Performance of the segmentation neural networks

We performed manual segmentation of the TRQ for the development dataset in accordance with the definition of the TRQ. Fig. 2a and 2b show an example of manual segmentation. All the segmented areas showed unimodal HU value distribution. The development dataset was divided into the training set and the test set. NN training and validation were performed using the training set.

**Fig. 2.**
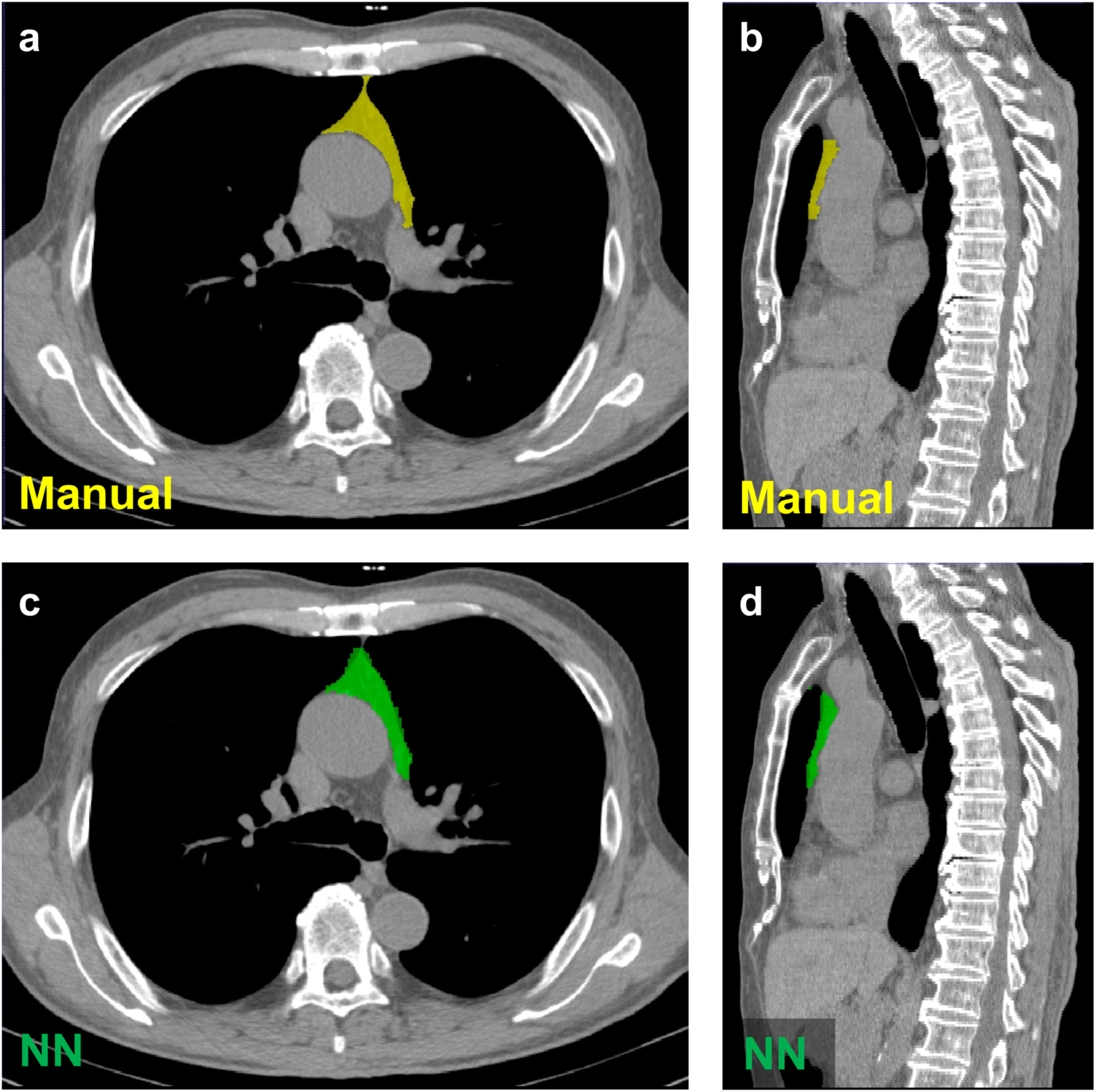
Example images of a TRQ segmented manually (**a** and **b**, the yellow region) and by a neural network (NN) model (**c** and **d**, the green region).

Automatic segmentation by NN models was conducted for the test set. Fig. 2c and 2d show an example of automatic segmentation by a NN. There were no invalid segmentation results. The median and interquartile range (IQR) of the DSC between the NN-segmented and manually segmented TRQ was 0.73 (IQR: 0.64-0.79) for the validation sets, and 0.76 (IQR: 0.67-0.82) for the test set. The Pearson correlation coefficients for the TRQ HU values and volumes were *r* = 0.996 and *r* = 0.986, and the ICCs were 0.996 and 0.985, respectively. Bland-Altman analysis revealed an acceptable level of bias for the TRQ HU values and volumes (mean difference: +0.43 HU and −0.14×10^4^ voxels, respectively). The yy plots and Bland-Altman plots are shown in Fig. 3.

**Fig. 3.**
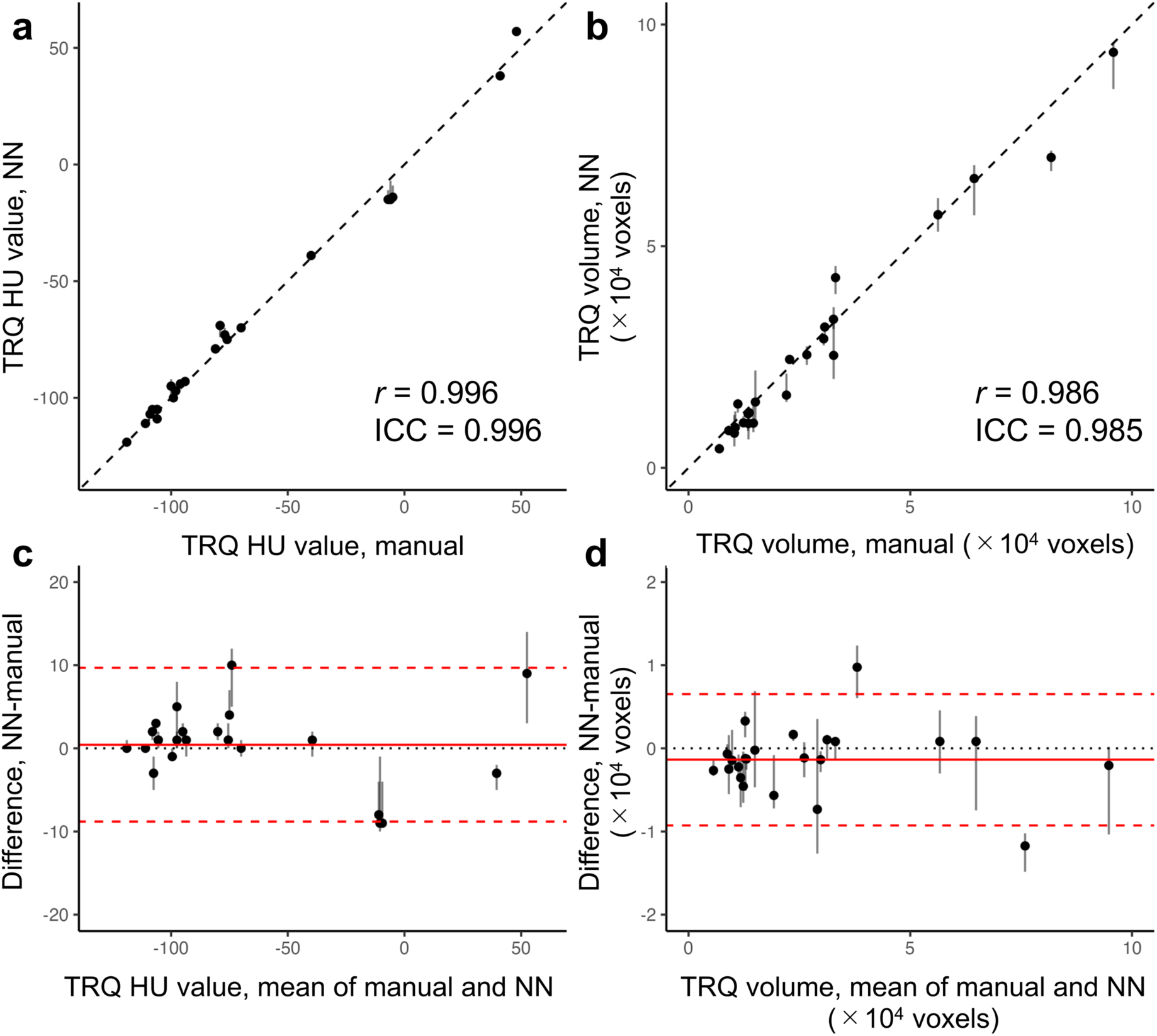
Evaluation of the TRQ segmentation quality. **a** and **b**, yy plots comparing the HU values and volumes obtained from the manually segmented and NN-segmented TRQs. *r* indicates the Pearson correlation coefficients. ICC indicates the intraclass correlation coefficients. The dashed lines are identity lines. **c** and **d**, Bland-Altman plots showing prediction errors of the TRQ HU values and volumes. The red lines indicate the mean difference, and the red dashed lines indicate the limits of agreement (LOA), calculated as mean±1.96× standard deviation. The error bars in the plots show the maximum and minimum values calculated from five TRQ segmentation results.

### Determination of the quality control criteria

Several measures correlated with the TRQ volume estimation error, HU value estimation error, and DSC (Fig. S3). The mean pairwise JS divergence, mean pairwise DSC, and HU value variance were the top three predictors of the TRQ volume estimation error and DSC. We selected these three as the quality control parameters. These parameters also predicted the TRQ HU value estimation error. In regard to the worst values of these parameters in the test set, we defined the quality control criteria as having been met when all of the following conditions were fulfilled: mean pairwise JS divergence < 0.1, mean pairwise DSC > 0.7, and HU value variance < 20. In the subsequent analyses, we excluded all the images that did not satisfy these criteria.

### Testing the theory for ETV calculation

The graphs and the regression analyses showed that the results were in complete agreement with the theoretical predictions (Supplementary Results, Fig. S4 and S5). This shows that our theory was applicable to actual data, and that the assumptions for calculation of the ETV were met. A strong correlation was found between the TRQ volume and TRQ HU value, underscoring the need to adjust for the effect of the adipose tissue volume. Based on the results, *A*_*adipose*_ = −110 HU was assumed for the subsequent analyses. *A*_*thymic*_ was assumed to be +80 HU, which is the reported approximate HU value of the newborn thymus [17].

### Age-related changes in the TRQ

The TRQ HU value, volume, and the ETV were calculated from 853 CT studies in the healthy dataset. There were no invalid segmentation results. In 30 CT studies (3.5 %), the automatic segmentation results failed to pass the quality control criteria. Age-related changes in the values are shown in Fig. 4 (*p_age_* < 0.001 for all). The TRQ HU value decreased with age and converged to around −110–-100 HU in the elderly. The TRQ volume increased with age, while the ETV decreased with age, which appeared mostly linear on a log scale. This suggests exponential decline of the thymic tissue volume with age. Female subjects showed significantly higher HU values and lower TRQ volumes (*p_sex_* < 0.001 for both), while no statistically significant difference in the ETV was noted between the male and female subjects (*p_sex_* = 0.19).

**Fig. 4.**
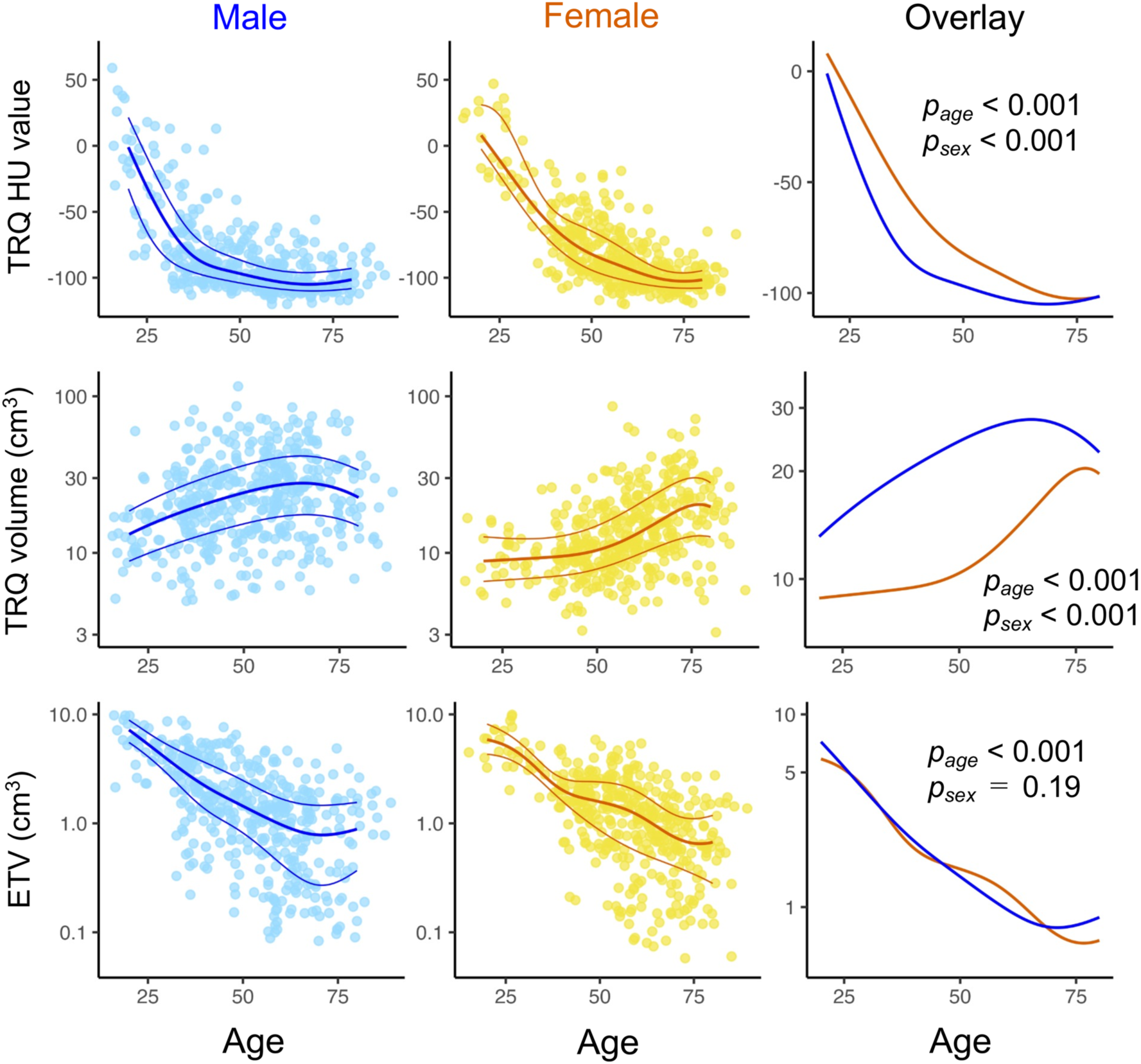
Age-related changes in the thymic region. Thick lines depict medians and thin lines depict interquartile ranges, estimated by quantile additive regression. The rightmost figures show an overlay of the median values in the male and female subjects. *p_age_* and *p_sex_* indicate the *p* values of the age and sex effects, respectively. TRQ volume and ETV are presented in log scale.

### ETV change in thymic hyperplasia

Among the 18 CT studies in the thymic hyperplasia dataset, one showed invalid segmentation results, and two failed to pass the quality control. The remaining CT studies with thymic hyperplasia showed ETV z-scores of significantly higher than zero (*p* < 0.001, Fig. 5a), suggesting that the ETV values in these subjects were higher as compared with those in age- and sex-matched healthy counterparts. The results of the ROC curve analysis, which yielded an AUC of 0.88 (95% CI: 0.75-1.0, Fig. 5b), showed that the ETV z-score could effectively differentiate between the thymic hyperplasia group and the healthy group. The cutoff value of the ETV z-score for this discrimination was estimated as 0.85 (sensitivity = 0.87, specificity = 0.86).

**Fig. 5.**
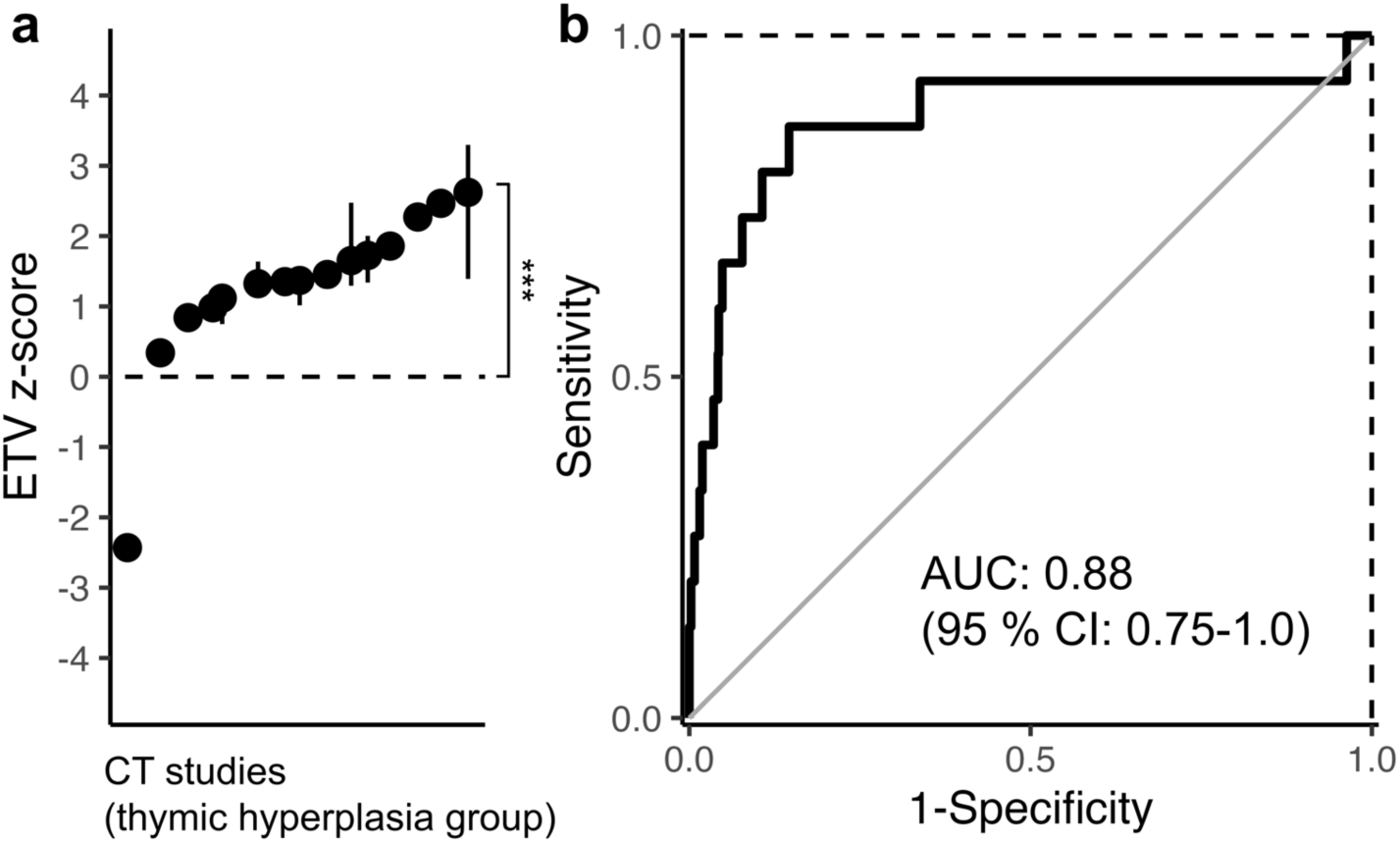
Evaluation of thymic hyperplasia using the ETV z-score. **a**, ETV z-scores of the thymic hyperplasia group, which were significantly higher than zero (\*\*\**p* < 0.001). The error bars indicate the maximum and minimum values. **b**, ROC curve analysis of the ETV z-score for differentiating between the group with thymic hyperplasia and the healthy group.

## Discussion

In this study, we could measure the volume and HU value of the thymic region with high robustness, using the neural networks. We established a quality control procedure to eliminate low-quality segmentations. Furthermore, we developed the ETV to estimate the thymic tissue volume. Our methodology could capture thymic tissue volume changes in age-related thymic involution and suspected thymic hyperplasia.

The segmentation quality as measured by DSC was not as high as the values reported for larger organs [18], reflecting the difficulty in automatic identification of the thymic region. Despite this, the volume and HU value of the NN-segmented TRQ showed a considerable degree of precision, indicating the effectiveness of our post-processing method.

ETV, a statistical estimator of the thymic tissue volume, was derived from a mathematical model based on simple assumptions. Although our theory ignored factors such as heterogeneity in the adipose and thymic tissues, the analyses strongly lent support to our theory in actual images, at least in healthy subjects. Furthermore, the ETV underwent an exponential decline with age. This is in line with the previously reported change of T-cell receptor excision circles (TREC), a biochemical indicator of thymic output, which also showed an exponential decline with age [19].

Whether the thymic tissue volume in adults differs between male and female subjects has been a matter of debate. Studies using HU value as a measure of thymic involution have mostly reported higher values in females [2, 10–12], while studies utilizing TREC [20–24] have reported a relatively small or non-significant difference. In line with this, we observed clearly higher HU value of the TRQ in female subjects. However, we did not observe a statistically significant difference in the ETV between male and female subjects. Conversely, the TRQ volume was significantly higher in male subjects. These data indicate that the sexual difference in the thymic region HU value is more dependent on the adipose tissue volume in the thymic region, rather than on the thymic tissue volume itself. Taken together with the significant effect of the TRQ volume on the HU value we observed, we think that the HU value alone cannot be used as a reliable indicator of thymic involution. The thymic visual scoring system may also not be appropriate, because it does not take the effect of adipose tissue volume into account.

Since no method existed to quantify the thymic tissue volume, the clinical significance of measuring the thymic tissue volume has not yet been established. Our thymic hyperplasia dataset contained several cases of autoimmune diseases such as rheumatoid arthritis and interstitial pneumonia. Our method might contribute to early detection of thymic hyperplasia and the underlying autoimmune disorders. It could also aid in evaluating the disease status and tracking the disease course. Further research is needed to explore these benefits of measuring/monitoring the thymic tissue volume.

This study had several important limitations. First, since homogeneity of the HU values within the TRQ is assumed, our method cannot evaluate heterogeneous changes in the thymic region, such as thymic tumors and nodular hyperplasia. Second, we could not obtain pathological evidence to determine how reliably the ETV predicted the actual thymic tissue volume. Importantly, since our thymic hyperplasia dataset did not include pathologically proven cases, we remain unable to make definitive statements regarding the diagnostic precision of our method. Third, our datasets included CT images acquired by different scanners and protocols, which could potentially have biased our results. Fourth, although we employed several strategies to control errors arising from mis-segmentation and obstacles, our data do not directly demonstrate their effectiveness.

In conclusion, our method enables robust quantification of thymic involution and hyperplasia, and our observations were consistent with previous reports. Our technique might prove useful in the management of diseases, including autoimmune disorders, which remains to be explored in future studies.

## Supporting information

Disclosure

## Data Availability

All data produced in the present study are available upon reasonable request to the authors.

