## Supplementary material for "Automated quantitative evaluation of thymic involution and hyperplasia on plain chest CT": Disclosure

1. ***Acknowledgements***

We thank Y. Itabashi, T. Inoue, H. Takahashi, and T. Seki for technical and ethical support. We also thank Dr. Sakiko Tsugawa for reviewing the manuscript. Lastly, we thank the study participants for their cooperation in this research.

2. ***Funding***

The authors state that this work has not received any funding.

***Compliance with Ethical Standards***

3. ***Guarantor:***

The scientific guarantor of this publication is Dr. Shin-Ichiro Kagami.

4. ***Conflict of Interest:***

Dr. Shin-Ichiro Kagami has received lecture fees from Janssen Pharmaceutical, AbbVie Inc, Nippon Boehringer Ingelheim, and VIATRIS. Dr. Kagami has also received a consulting fee from Asahi Kasei Pharma.

5. ***Statistics and Biometry:***

One of the authors has significant statistical expertise.

6. ***Informed Consent:***

Written informed consent was waived by the Institutional Review Board.

7. ***Ethical Approval:***

Institutional Review Board approval was obtained.

8. ***Study subjects or cohorts overlap:***

No study subjects or cohorts have been previously reported.

9. ***Methodology***

Methodology:

- retrospective
- observational
- performed at one institution
