## Supplementary Material for "Automated quantitative evaluation of thymic involution and hyperplasia on plain chest CT"

#### Supplementary Methods

##### Image acquisition and data conversion

All computed tomography (CT) images were acquired using one of the following multi-detector CT scanners: SOMATOM Emotion 16 (Siemens), SOMATOM Sensation Cardiac (Siemens), SOMATOM Definition AS (Siemens), SOMATOM Emotion 6 (Siemens), Revolution Frontier (GE HealthCare), Aquilion (Canon Medical Systems), or Aquilion ONE (Canon Medical Systems). The CT images were extracted as DICOM files and converted to the NIfTI format using dcm2niix [1]. The ITK-SNAP software [2] was used for visual inspection of the CT images. Radiological parameters were obtained from the dcm2niix output.

##### Training of segmentation neural networks

For automatic identification of the thymic region for quantification (TRQ) in CT images, we trained DeepLabV3 neural network (NN) models with the ResNet-50 backbone [3]. The DeepLabV3 NN is known to show high performance in semantic segmentation tasks. The NN models were implemented using Python (v3.9) and PyTorch (v1.10).

Each CT image was processed by the neural network model as follows (see Supplementary Fig. 1). Each axial slice image was used as an input to the NN. The original $512 \times 512$ slice images were downscaled to the size of $256 \times 256$ before being inputted. The NN outputted label information in $256 \times 256$. The labels "1", "2", and "0" were assigned to the airways, the TRQ, and none of these, respectively. The output was upscaled to $512 \times 512$ for the subsequent analyses.

The development dataset was split randomly into six subsets, stratified by the age, sex, and slice thickness using the iterative-stratification package [4]. One subset was used as the test set, while the remaining five subsets (training set) were used for NN training and five-fold cross-validation. During the five-fold cross-validation process, we optimized the conditions for image augmentation and performed hyperparameter tuning.

During the training process, the following image augmentation processing was performed: Rotation: The slice image was rotated by a random angle from -30° to +30°; Random erasing: Up to three rectangular regions of random sizes were erased, and the inner areas were replaced with random values. The parameter settings for the NN training were as follows: batch size: 8; number of epochs: 20; learning rate: 0.0001. RMSprop was used as the optimization algorithm. The loss function was cross entropy.

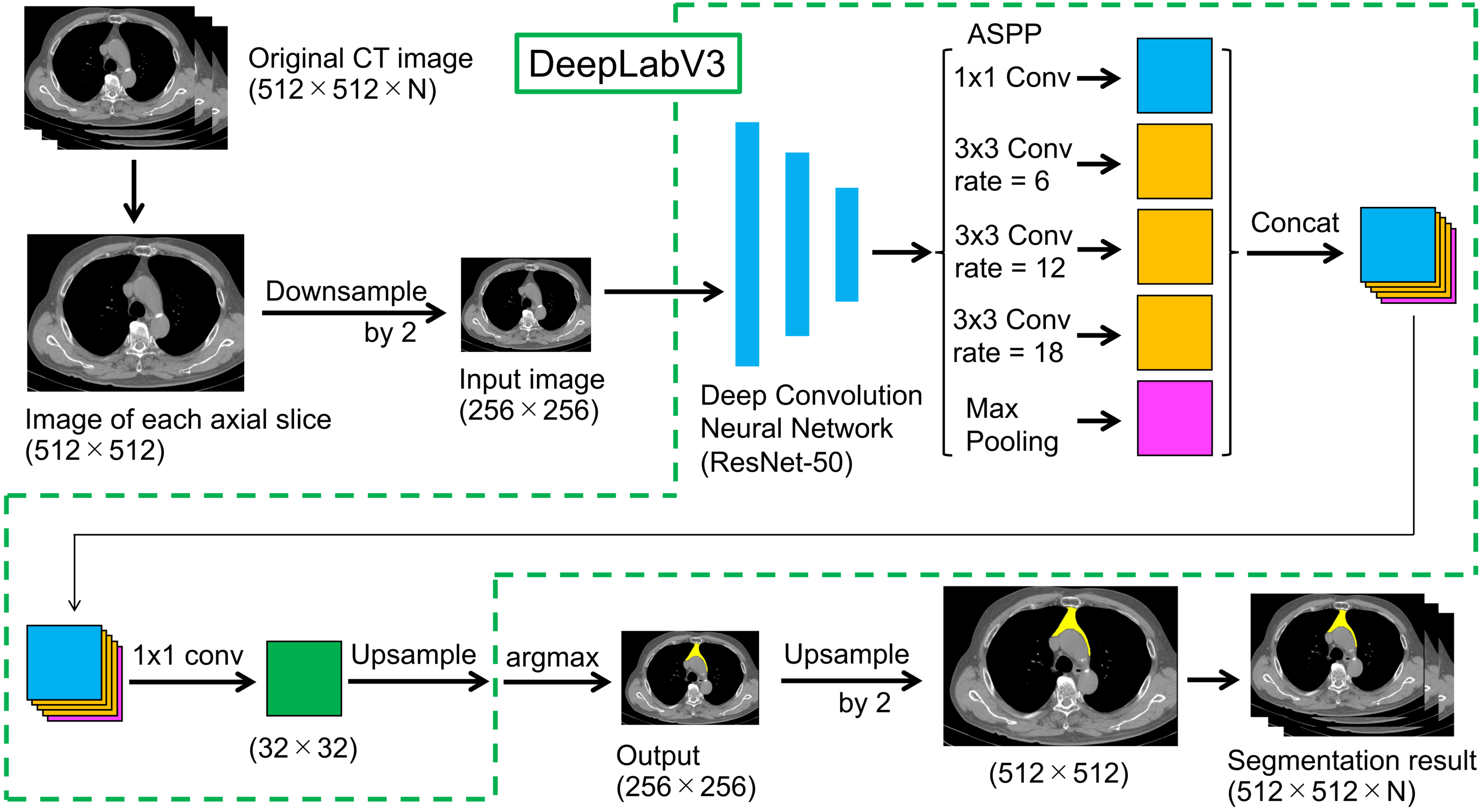


**Fig. S1** Schematic illustration of the automatic segmentation process by the neural network. The DeepLabV3 neural network architecture utilizes the atrous spatial pyramid pooling (ASPP) to integrate the image information at multiple scales. The numbers in the parentheses indicate the image or tensor sizes.

##### Measurement of the TRQ HU value

The TRQ in each CT study was segmented by the five NN models generated in the five-fold cross-validation. For robust measurement of the TRQ HU value in the presence of potential mis-segmentation and obstacles, the five segmentation results were separately processed as follows: First, the distribution of the HU value in the segmented TRQ was obtained. Next, the density function was estimated using kernel density distribution estimation (KDE). KDE was estimated by the scipy.stats.gaussian_kde function in the SciPy package [5]. Finally, the mode value, where the density function reaches its maximum, was set as the representative HU value of the segmented TRQ. The mode value is robust against outliers arising from mis-segmentation and obstacles. It is an appropriate central tendency to use when the distribution is unimodal. When the probability density function in a segmented TRQ was multimodal, the segmentation result was labeled as invalid and discarded; we considered probability distributions as multimodal when the second highest peak of the density function exceeded half the height of the highest peak.

##### Establishing the quality control criteria

The segmentation quality was evaluated by three indicators: TRQ volume estimation error (absolute log fold-difference between the NN-segmented and manually segmented TRQ volume), TRQ HU value estimation error (absolute difference between the NN-segmented and manually segmented TRQ HU value), and DSC between the NN-segmented and manually segmented TRQ. To predict these indicator values, seven measures were calculated from the five automatic segmentation results in each study as follows.

1. HU value median: Median of the mode HU values calculated for each segmentation result.
2. Volume median: Median of the TRQ volumes calculated for each segmentation result.
3. HU value variance: Unbiased variance of the mode HU values calculated for each segmentation result.
4. Volume CV: Coefficients of variation (CV) of the TRQ volumes calculated for each segmentation result. CV is the square root of unbiased variance divided by the mean.
5. Mean pairwise JS divergence: Jensen-Shannon divergence (JS divergence) evaluates the dissimilarity between two probability distributions. JS divergence of the HU value distributions was calculated between all possible pairs and averaged. To estimate the probability distribution, the HU values were limited to the range falling between -300 HU and 300 HU and smoothed by KDE.
6. Mean pairwise DSC: DSC was calculated for all possible pairs and averaged.
7. HU value second peak height: The height of the second highest peak divided by the height of the highest peak in HU value distribution was calculated for each segmented TRQ and averaged. The distribution was estimated by KDE. When the second highest peak could not be detected, the value was calculated as zero.

The correlations between the three quality indicators and seven measures were evaluated by pairwise Spearman rank correlation in the test set. The $p$ values were FDR-adjusted by the Benjamini-Hochberg method. Multivariable analyses were not performed due to the limited sample size.

**Derivation of the estimator of thymic volume (ETV)**

When the TRQ is considered as a homogenous mixture of thymic and adipose tissue, and it is assumed that the volumes of the thymic and adipose tissue are $v_{thymic}$ and $v_{adipose}$ with constant HU values of $A_{thymic}$ and $A_{adipose}$ ($A_{thymic} > A_{adipose}$), respectively, TRQ volume $v_{TRQ}$ and HU value $A_{TRQ}$ would have the following relationships:

$$v_{TRQ} = v_{thymic} + v_{adipose}$$

and

$$A_{TRQ} = \frac{v_{thymic}A_{thymic} + v_{adipose}A_{adipose}}{v_{thymic} + v_{adipose}} .$$

From these equations, we obtain

$$(A_{TRQ} - A_{adipose})v_{TRQ} = (A_{thymic} - A_{adipose})v_{thymic} \tag{1}$$

and

$$\log_{10}(A_{TRQ} - A_{adipose}) = -\log_{10} v_{TRQ} + \log_{10} v_{thymic} - \log_{10}(A_{thymic} - A_{adipose}) . \tag{2}$$

In the actual data analysis, $A_{TRQ}$ and $v_{TRQ}$ were estimated from the segmentation results. We denote these as $\widehat{A_{TRQ}}_i$ and $\widehat{v_{TRQ}}_i$ for the CT study $i$, respectively.

When the values of the constants $A_{thymic}$ and $A_{adipose}$ are correctly assumed and thymic tissue volume $v_{thymic_i}$ is also considered as a constant, $\widehat{A_{TRQ}}_i$ and $\widehat{v_{TRQ}}_i$ should satisfy the equations

$$\left(\widehat{A_{TRQ}}_i - A_{adipose}\right)\widehat{v_{TRQ}}_i = C \tag{1$'$}$$

and

$$\log_{10}\left(\widehat{A_{TRQ}}_i - A_{adipose}\right) = -\log_{10}\widehat{v_{TRQ}}_i + C' \tag{2$'$}$$

where $C$ and $C'$ are constant. It can be inferred from formula $(1)'$, that when $\widehat{v_{TRQ}}_i$ approaches infinity, $\widehat{A_{TRQ}}_i$ converges to $A_{adipose}$. Also, regarding formula $(2)'$, the regression model

$$\log_{10}\left(\widehat{A_{TRQ}}_i - A_{adipose}\right) = \beta_1 \log_{10}\widehat{v_{TRQ}}_i + \beta_0 + \varepsilon_i , \tag{2$''$}$$

where $\beta_1$ and $\beta_0$ are regression coefficients, is expected to bear a result equivalent to $\beta_1 = -1$. $\varepsilon_i$ is the error term.

We sought to test whether these theories would hold true for actual data. Using the healthy dataset, the relationship between $\widehat{A_{TRQ}}_i$ and $\widehat{v_{TRQ}}_i$ was assessed using a scatterplot and approximation curves to see if equations $(1)'$ and $(2)'$ hold true. The curves were drawn using the geom_smooth function in the ggplot package with default options. Since $v_{thymic_i}$ was assumed to be constant when deriving equations $(1)'$ and $(2)'$, data were grouped based on the age when drawing the curves, expecting $v_{thymic_i}$ to be roughly constant within each age group. Furthermore, regression model $(2)''$ was applied to the healthy dataset as a generalized linear mixed model. Since the regression model also assumes that $v_{thymic_i}$ is constant across images, $\beta_0$ was set as a random effect dependent on the age group. Fixed effect $\beta_1$ was estimated assuming different values of $A_{adipose}$. $\widehat{A_{TRQ}}_i$ values smaller than $A_{adipose}$ were removed, or adjusted as

$$\widehat{A'_{TRQ}}_i = \begin{cases} \widehat{A_{TRQ}}_i, \text{ if } \widehat{A_{TRQ}}_i \geq A_{adipose} + \delta_A \\ A_{adipose} + \delta_A, \text{ if } \widehat{A_{TRQ}}_i < A_{adipose} + \delta_A \end{cases} \quad (3)$$

where $\delta_A$ is the constant of margin. $\delta_A = 1\ \text{HU}$ was used for all the analyses. The lme4 package [6], lmertest package [7] and performance package [8] were used for the generalized linear mixed model analyses.

Finally, given that the assumptions made above are true, $v_{thymic_i}$ for each CT image can be estimated from $\widehat{A_{TRQ}}_i$ and $\widehat{v_{TRQ}}_i$ by an estimator derived from equation $(1)$, defined as

$$\text{ETV}\left(\widehat{A'_{TRQ}}_i, \widehat{v_{TRQ}}_i\right) \stackrel{\text{def}}{=} \frac{\widehat{A'_{TRQ}}_i - A_{adipose}}{A_{thymic} - A_{adipose}} \widehat{v_{TRQ}}_i.$$

This is the estimator of thymic volume (ETV), a statistical estimator of the thymic tissue volume. $\widehat{A_{TRQ}}_i$ was adjusted as in formula $(3)$. As we assumed arbitrary constant values for $A_{adipose}$ and $A_{thymic}$, ETV is a relative (not absolute) measure of the actual thymic tissue volume.

##### Measuring age-related changes of the thymic region

Due to the nonlinear and highly variable nature of the aging process of the thymus, our data were expected to show nonlinearity and heteroskedasticity. Hence, we utilized quantile additive regression models to analyze the data. To reveal age-related changes in the median and the interquartile range of the TRQ HU value, volume, and ETV, we applied the following quantile additive regression model to the healthy dataset, separately for males and females:

$$y = f_{age}(\text{age}_i) + \varepsilon_i\ .$$

$y$ denotes the TRQ HU value, volume, or ETV. $f(\cdot)$ and $\varepsilon_i$ indicate smooth terms and error terms, respectively. $f_{age}(\text{age}_i)$ represents the effect of age. Alternatively, to test for the effect of sex, the following model was applied:

$$y = f_{age}(\text{age}_i) + \text{sex}_i f_{sex}(\text{age}_i) + \varepsilon_i\ ,$$

where the term $\text{sex}_i f_{sex}(\text{age}_i)$ represents the age-dependent and age-independent effects of sex. $p$ values for $f_{age}(\text{age}_i)$ and $\text{sex}_i f_{sex}(\text{age}_i)$ are reported as $p_{age}$ and $p_{sex}$ , respectively, for median values. The qgam package [9] was used for all of these quantile additive regression analyses. Options k=10 and bs="ad" were used in the s function.

##### Calculation of the ETV z-score

To make the ETV value understandable in comparison with the healthy group, the ETV z-score was defined as a robust z-score [10] calculated from the common logarithm of the ETV value, as

$$\begin{aligned} \text{ETV z-score}_i &\stackrel{\text{def}}{=} \frac{\log_{10}\text{ETV}_i - Q_{\log_{10}\text{ETV}}(0.5;\ \text{age}_i, \text{sex}_i)}{0.74 \times \left(Q_{\log_{10}\text{ETV}}(0.75;\ \text{age}_i, \text{sex}_i) - Q_{\log_{10}\text{ETV}}(0.25;\ \text{age}_i, \text{sex}_i)\right)} \\ &= \frac{\log_{10}\text{ETV}_i - \log_{10} Q_{\text{ETV}}(0.5;\ \text{age}_i, \text{sex}_i)}{0.74 \times (\log_{10} Q_{\text{ETV}}(0.75;\ \text{age}_i, \text{sex}_i) - \log_{10} Q_{\text{ETV}}(0.25;\ \text{age}_i, \text{sex}_i))}, \end{aligned}$$

where $Q_{\log_{10}\text{ETV}}(p;\ \text{age}_i, \text{sex}_i)$ and $Q_{\text{ETV}}(p;\ \text{age}_i, \text{sex}_i)$ represent the quantile functions of $\log_{10}\text{ETV}_i$ and $\text{ETV}_i$ respectively. $p = 0.25, 0.5, 0.75$ correspond to the quartile points. Values of $Q_{\text{ETV}}(p;\ \text{age}_i, \text{sex}_i)$ were calculated from the quantile regression models applied to the healthy dataset. Log transformation was performed because the ETV had right-skewed distribution.

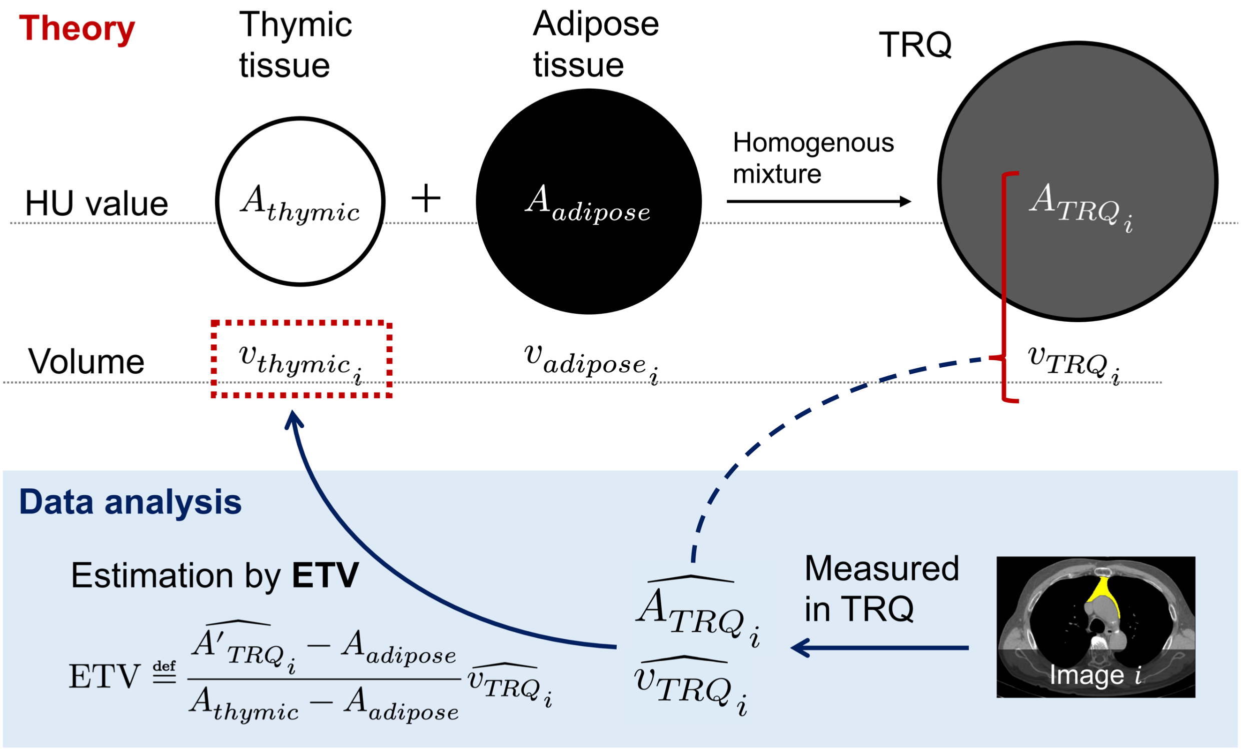


**Fig. S2** Diagram depicting the theory and the calculation process of the estimator of thymic volume (ETV). The statistical estimator ETV estimates the thymic tissue volume based on the volume and HU value of the segmented TRQ, in a way similar to solving mixture problems in elementary school math.

### Supplementary Results

#### Validation of the ETV theory

The TRQ HU value showed a negative correlation with the TRQ volume (Fig. S4a and S4b). The graphs showed hyperbola-like shapes across age groups, as predicted by equation $(1)'$. The HU value converged to around -110 HU when the TRQ volume was large, which suggests the true value of $A_{adipose}$. On the log-log plot, the relationship was roughly linear with slope -1 across age groups (Fig. S4c), in agreement with formula $(2)'$. Curves from older groups appeared in the lower positions, reflecting thymic involution with age (Fig. 4b and 4c).

In the regression analyses, $\beta_1$ mostly matched the predicted value -1 when $A_{adipose}$ was assumed to be $-110$ HU. Effect of the TRQ volume on the TRQ HU value was strong and statistically significant in the model assuming $A_{adipose} = -110$ HU ($p < 0.001$, marginal $R^2 = 0.30$).

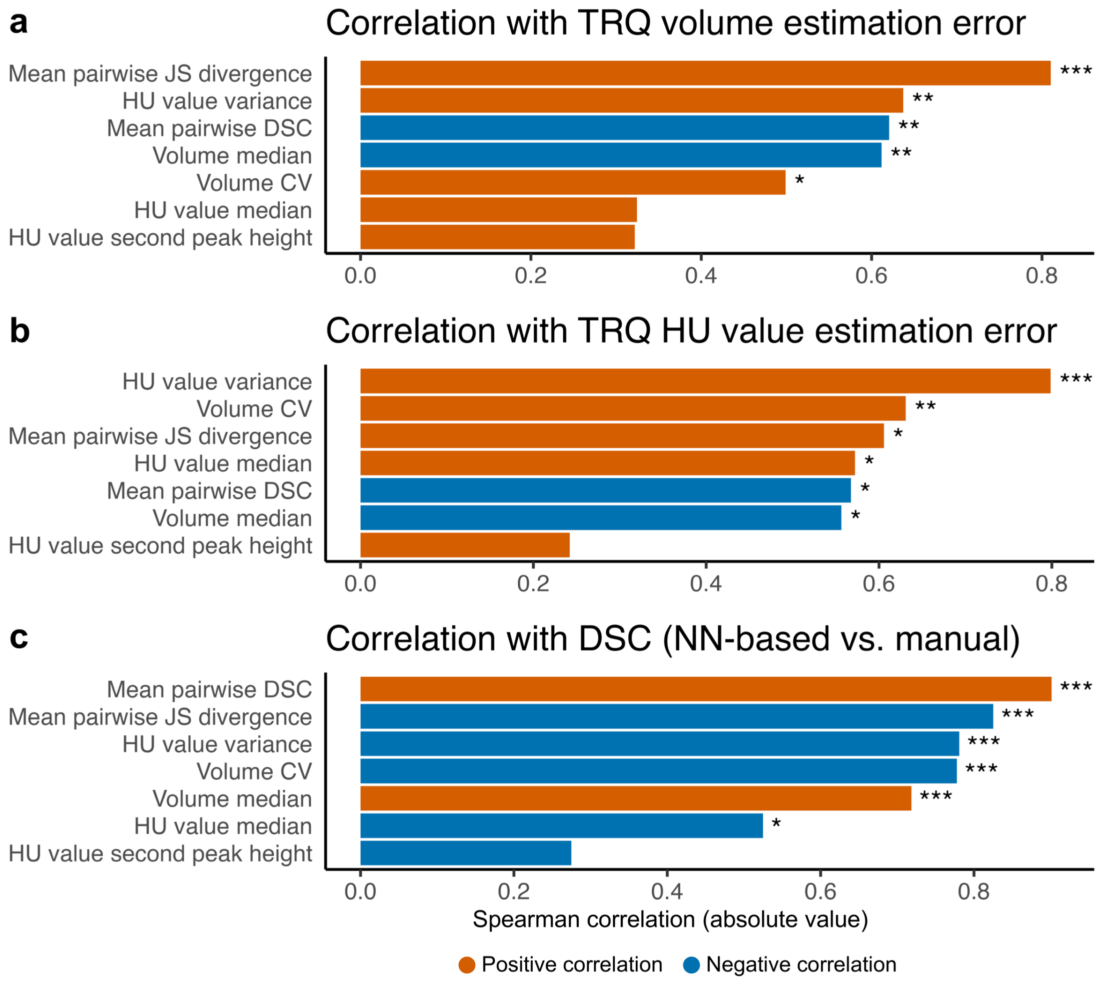


**Fig. S3** Results of the correlation analyses to determine the quality control criteria. Correlations between seven measures and the segmentation quality indicators were calculated. The quality indicators were the TRQ volume estimation error (**a**), HU value estimation error (**b**), and DSC (NN-based vs. manual) (**c**). *$p < 0.05$, **$p < 0.01$, ***$p < 0.001$.

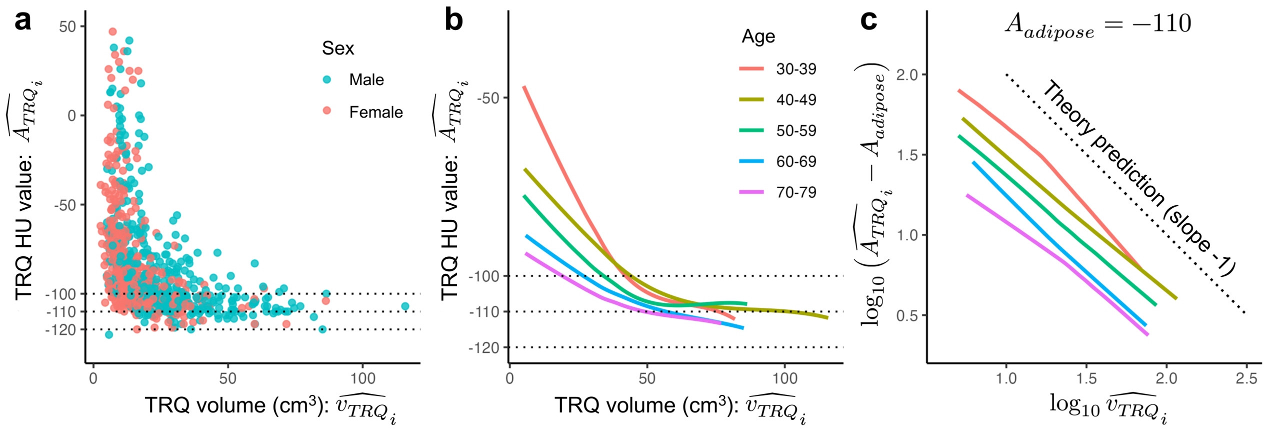


**Fig. S4** Relationship between the TRQ HU value and TRQ volume. **a** and **b**, The relationship is shown on a linear scale, on a scatterplot (**a**) and spline curves (**b**). The data points were grouped based on age when drawing the curves. **c**, The relationship is shown on a log-log scale. $A_{adipose}$ was assumed to be -110 HU. The dotted line shows slope -1 for reference.

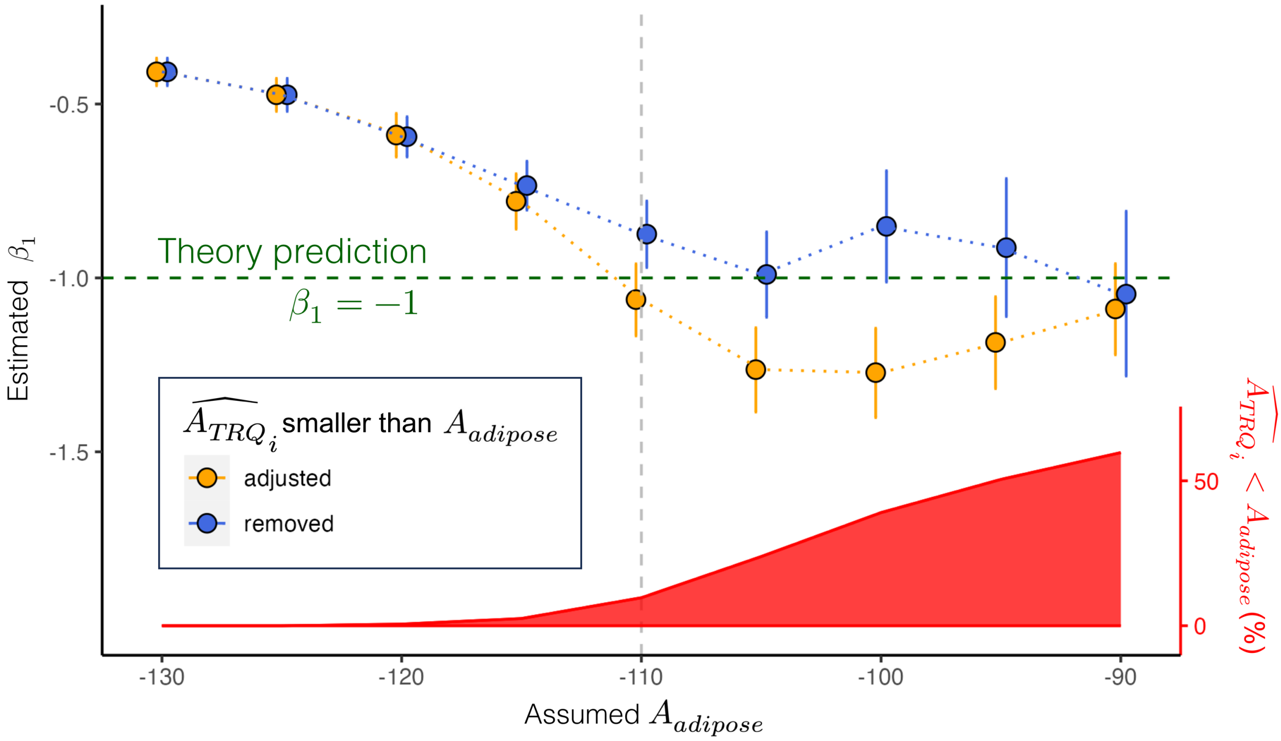


**Fig. S5** Results of the regression analyses using model $(2)''$. $\beta_1$ estimated for different values of $A_{adipose}$ are shown as blue and orange points. Studies with TRQ HU value smaller than $A_{adipose}$ were either adjusted (orange) or removed (blue) from the analyses. The error bars depict the confidence intervals. The red ribbon graph at the bottom shows the percentage of studies with TRQ HU values smaller than the assumed $A_{adipose}$.
